# Analysis of blood and nasal epithelial transcriptomes to identify mechanisms associated with control of SARS-CoV-2 viral load in the upper respiratory tract

**DOI:** 10.1101/2023.03.09.23287028

**Authors:** Mahdi Moradi Marjaneh, Joseph D Challenger, Antonio Salas, Alberto Gómez-Carballa, Abilash Sivananthan, Irene Rivero-Calle, Gema Barbeito-Castiñeiras, Cher Y Foo, Yue Wu, Felicity Liew, Heather R Jackson, Dominic Habgood-Coote, Giselle D’Souza, Samuel Nichols, Victoria J Wright, Michael Levin, Myrsini Kaforou, Ryan S Thwaites, Lucy C Okell, Federico Martinón-Torres, Aubrey J Cunnington, PERFORM Consortium, GEN-COVID Study Group

**Affiliations:** Section of Paediatric Infectious Disease, Department of Infectious Disease, Imperial College London, London, UK; Centre for Paediatrics and Child Health, Imperial College London, London, UK; Section of Virology, Department of Infectious Diseases, Faculty of Medicine, Imperial College London, London, UK; Medical Research Council Centre for Global Infections Disease Analysis, Department of Infectious Disease Epidemiology, Imperial College London, London, UK; Unidade de Xenética, Instituto de Ciencias Forenses, Facultade de Medicina, Universidade de Santiago de Compostela, and GenPoB Research Group, Instituto de Investigación Sanitaria (IDIS), Hospital Clínico Universitario de Santiago (SERGAS), Galicia, Spain; Genetics, Vaccines and Infections Research Group (GENVIP), Instituto de Investigación Sanitaria de Santiago, Universidade de Santiago de Compostela, Santiago de Compostela, Galicia, Spain; Centro de Investigación Biomédica en Red de Enfermedades Respiratorias (CIBER-ES), Madrid, Spain; Translational Pediatrics and Infectious Diseases, Department of Pediatrics, Hospital Clínico Universitario de Santiago de Compostela, Santiago de Compostela, Galicia, Spain; Servicio de Microbiología y Parasitología, Complejo Hospitalario Universitario de Santiago de Compostela, Santiago de Compostela, Galicia, Spain; School of Medicine, Imperial College London, London, UK; Department of Surgery and Cancer, Imperial College London, St. Mary’s Hospital, London, UK; National Heart and Lung Institute, Imperial College London, London, UK

**Keywords:** COVID-19, SARS-CoV-2, upper respiratory tract, viral load, mathematical modelling, transcriptome, gene network analysis

## Abstract

**Background:** The amount of SARS-CoV-2 detected in the upper respiratory tract (URT viral load) is a key driver of transmission of infection. Current evidence suggests that mechanisms constraining URT viral load are different from those controlling lower respiratory tract viral load and disease severity. Understanding such mechanisms may help to develop treatments and vaccine strategies to reduce transmission. Combining mathematical modelling of URT viral load dynamics with transcriptome analyses we aimed to identify mechanisms controlling URT viral load.

**Methods:** COVID-19 patients were recruited in Spain during the first wave of the pandemic. RNA sequencing of peripheral blood and targeted NanoString *n*Counter transcriptome analysis of nasal epithelium were performed and gene expression analysed in relation to paired URT viral load samples collected within 15 days of symptom onset. Proportions of major immune cells in blood were estimated from transcriptional data using computational differential estimation. Weighted correlation network analysis (adjusted for cell proportions) and fixed transcriptional repertoire analysis were used to identify associations with URT viral load, quantified as standard deviations (z-scores) from an expected trajectory over time.

**Results:** Eighty-two subjects (50% female, median age 54 years (range 3-73)) with COVID-19 were recruited. Paired URT viral load samples were available for 16 blood transcriptome samples, and 17 respiratory epithelial transcriptome samples. Natural Killer (NK) cells were the only blood cell type significantly correlated with URT viral load z-scores (r = -0.62, *P* = 0.010). Twenty-four blood gene expression modules were significantly correlated with URT viral load z-score, the most significant being a module of genes connected around *IFNA14* (Interferon Alpha-14) expression (r = -0.60, *P* = 1e-10). In fixed repertoire analysis, prostanoid-related gene expression was significantly associated with higher viral load. In nasal epithelium, only *GNLY* (granulysin) gene expression showed significant negative correlation with viral load.

**Conclusions:** Correlations between the transcriptional host response and inter-individual variations in SARS-CoV-2 URT viral load, revealed many molecular mechanisms plausibly favouring or constraining viral load. Existing evidence corroborates many of these mechanisms, including likely roles for NK cells, granulysin, prostanoids and interferon alpha-14. Inhibition of prostanoid production, and administration of interferon alpha-14 may be attractive transmission-blocking interventions.

## Background

The advent of Severe Acute Respiratory Syndrome Coronavirus 2 (SARS-CoV-2) leading to the coronavirus disease 2019 (COVID-19) has placed an enormous burden on affected individuals, healthcare systems, and economies worldwide. SARS-CoV-2 is highly transmissible and causes a wide range of severity from asymptomatic infection to severe disease and death. The amount of SARS-CoV-2 detected in the upper respiratory tract of infected individuals (URT viral load) is a key driver of transmission of infection (1). High URT viral loads can increase household and non-household transmissions by up to nearly 60% and 40%, respectively (2). Interestingly, URT viral load does not necessarily correlate with severity of illness, nor is it determined by established risk factors for poor outcome such as age and sex (3, 4). This suggests that the host immune mechanisms involved in constraining the virus in the URT are different from those determining the severity of illness, although such mechanisms have not been fully elucidated. In contrast, high and persistent SARS-CoV-2 shedding in the lower respiratory tract (LRT) is associated with severe disease (5), indicating differences in the mechanisms underlying control and pathogenesis of SARS-CoV-2 in the URT and LRT. Understanding the mechanisms controlling the viral load in the URT could illuminate new strategies to prevent transmission from infected individuals and might also enable control of the localised infection before it progresses to the LRT, triggering more serious illness.

URT viral load is highly dynamic. It changes over the course of illness due to dynamic interactions with the host immune response; it peaks around the time of symptom onset and then gradually decreases to a low level over the following 10 days (6, 7). Moreover, the kinetics of viral load vary between individuals, presumably determined by variation in immune responses (3). The host response constraining viral load includes both an immediate innate component and a later adaptive response (3, 8, 9). With limited *in vivo* data, researchers have attempted to mathematically model and explain the viral-host interaction and host immune responses to better understand the dynamics of SARS-CoV-2 viral load. We have recently developed a within-host model that has been successful in interpreting URT viral load kinetics in a wide range of data including 2172 serial measurements from 605 subjects, collected from 17 different studies (3).

Despite the dynamic interaction between the virus and host immune system during SARS-CoV-2 infection and the diversity in such interaction observed between individuals, the immune response involves conserved elements which can be reflected in host transcriptomes (10). While gene expression is a dynamic process, and a single transcriptomic experiment usually captures only a “snapshot” in time, using robust transcriptional analyses we can pinpoint key biological mechanisms underlying the immune response. The host transcriptomic response in human infection is often studied in peripheral blood leukocytes. This is because peripheral blood leukocytes mount cell-intrinsic responses to pathogens but also mount transcriptional responses to signals arising from the organs through which they circulate. Evaluating the host transcriptome in the context of the dynamics of host-pathogen interaction can be a powerful approach to elucidate mechanisms responsible for control of pathogen load (11).

Here we sought to combine mathematical modelling of URT SARS-CoV-2 viral load dynamics in individual subjects with the analysis of peripheral blood and nasal epithelium transcriptomes to identify mechanisms associated with the control of viral load. The mechanistic correlates of URT viral load identified herein may be important to develop new therapeutic and vaccine strategies to block transmission of SARS-CoV-2.

## Results

### Participants

We performed transcriptome analyses for 82 COVID-19 patients (50% female, median age 54 years (range 3-73 years)) recruited during the “first wave” of COVID-19 in Spain, before vaccination and natural infection became determinants of the immune response to SARS-CoV-2. Whole blood RNA sequencing (RNA-Seq) and nasal epithelium *n*Counter NanoString gene expression assay data were generated (see Methods) for 68 and 24 subjects, respectively, with 10 subjects being included in both analyses. Clinical characteristics of all subjects are provided in **Supplementary Table 1**.

The whole blood transcriptome profiles were used to construct gene co-expression networks and detect clusters of interconnected genes (see below). For gene module discovery, to optimise the generalisability of modules, we included all 68 COVID-19 subjects with RNA-Seq data (regardless of whether they had co-infections), and an additional 18 uninfected healthy control subjects and 9 subjects with non-COVID-19 infections (4 bacterial and 5 viral) (**Supplementary Table 1)**, all sequenced in the same batch. However, of the COVID-19 cases who were free from suspected or proven bacterial co-infections, only 16 had URT viral load measurement and RNA samples collected on the same day and within 15 days of symptom onset (a time window during which the replicating virus can be isolated (12)). Only these 16 subjects were included in analyses correlating URT viral load with the whole blood transcriptome (**Table 1**, **Figure 1A**). Subjects were mostly female (57.1%), with ages ranging from 3 to 78 years (median = 55 years) (**Figures 1B and 1C**). The disease severity was mild (n = 3; 19%), moderate (n = 7; 44%), and severe (n = 6; 37%).

**Figure 1.**
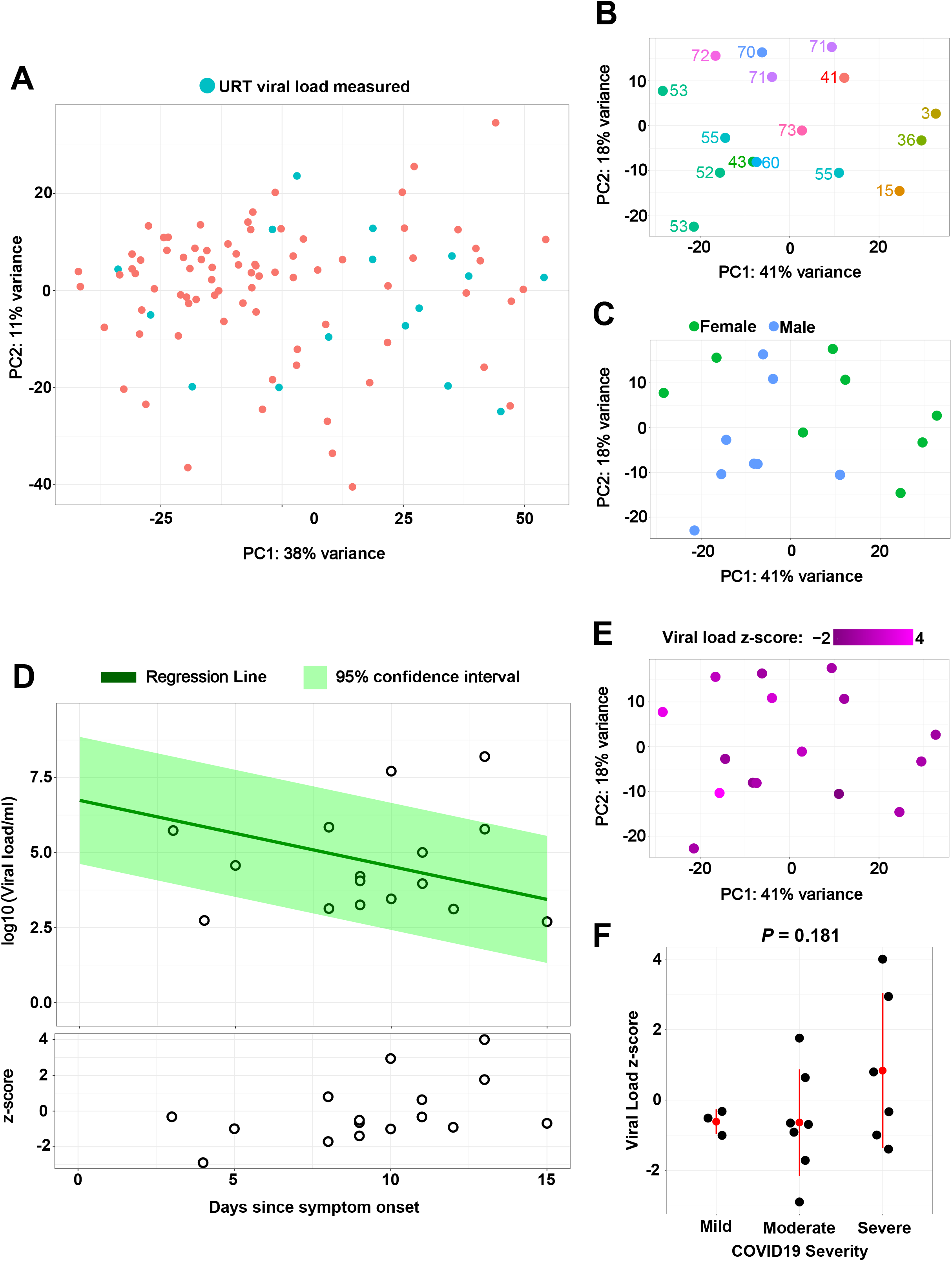
Overview of peripheral blood gene expression and viral load in subjects with COVID-19. A) PCA (principal component analysis) plot of peripheral blood gene expression determined by RNA-Seq. Samples with paired URT viral load measurement are coloured as blue. B) and C) PCA plots represent samples with paired RNA-Seq and viral load data coloured by age and sex, respectively. D) Calculation of viral load z-scores. In the upper panel, the viral load data of the present study (black circles) are plotted against the time since symptom onset. The green line indicates a linear regression model fitted to the viral load data from 16 different datasets previously studied. The shaded green area represents the 95% confidence interval for the regression model. As shown in the lower panel, for each data point, a z-score is calculated as the distance of the data point from the mean trajectory (green line). E) PCA plot of samples with paired data coloured based on viral load z-score. F) Viral load z-score is compared between groups of different COVID-19 severity. Red dots and whiskers represent mean and 1 standard deviation.

**Table 1.** Samples used to correlate URT viral load and whole blood transcriptome

| Days of illness<sup>1</sup> | Age range (year) | Sex | Severity | Average Ct value<sup>2</sup> | Calculated viral load (log10 (viral load/ml)) | Viral load z-score |
| --- | --- | --- | --- | --- | --- | --- |
| 3 | 11-15 | Female | Mild | 27.72 | 5.49 | -0.32 |
| 4 | 51-55 | Male | Moderate | 37.08 | 2.52 | -2.89 |
| 5 | 66-70 | Male | Severe | 31.53 | 4.34 | -0.99 |
| 8 | 71-75 | Female | Severe | 35.87 | 5.60 | 0.80 |
| 8 | 41-45 | Male | Moderate | 27.41 | 2.91 | -1.71 |
| 9 | 51-55 | Male | Severe | 35.43 | 3.04 | -1.39 |
| 9* | 51-55 | Male | Moderate | 32.36 | 3.83 | -0.65 |
| 9 | 36-40 | Female | Mild | 33.1 | 3.98 | -0.51 |
| 10 | 1-5 | Female | Mild | 21.03 | 3.23 | -1.00 |
| 10 | 51-55 | Female | Severe | 34.39 | 7.45 | 2.94 |
| 11 | 71-75 | Female | Moderate | 29.97 | 4.77 | 0.64 |
| 11 | 56-60 | Male | Severe | 33.24 | 3.73 | -0.33 |
| 12 | 41-45 | Female | Moderate | 36.64 | 2.90 | -0.91 |
| 13* | 51-55 | Male | Severe | 28.62 | 7.93 | 4 |
| 13 | 71-75 | Male | Moderate | 19.57 | 5.54 | 1.76 |
| 15 | 71-75 | Female | Moderate | 37.17 | 2.48 | -0.69 |

<sup>1</sup> How many days after symptom onset the viral load was measured.
<sup>2</sup> Cycle threshold value
\* These two samples are from the same subject

We also performed a *n*Counter NanoString gene expression analysis on nasal epithelium samples from 24 COVID-19 patients, including 17 with URT viral load measurement within 15 days from symptom onset (**Table 2**). The subjects’ ages ranged from 16 to 80 years (median = 47 years), and most had mild disease (n = 9; 53%) or severe disease (n = 6; 35.3%).

**Table 2.** Subjects used to correlate URT viral load with nasal epithelium NanoString profiles

| Days of illness | Age range (year) | Sex | Severity | Average Ct value | Calculated viral load (log10 (viral load/ml)) | Viral load z-score |
| --- | --- | --- | --- | --- | --- | --- |
| 0 | 26-30 | Female | Mild | 27.53 | 5.94 | -0.74 |
| 3 | 16-20 | Female | Mild | 27.72 | 5.74 | -0.32 |
| 4 | 76-80 | Female | Severe | 25.36 | 6.46 | 0.56 |
| 5 | 66-70 | Male | Severe | 31.53 | 4.57 | -0.99 |
| 6 | 41-45 | Male | Mild | 38.16 | 2.39 | -2.8 |
| 7 | 46-50 | Male | Mild | 29.9 | 5.01 | -0.17 |
| 8 | 71-75 | Female | Severe | 27.41 | 5.85 | 0.8 |
| 8 | 31-35 | Female | Mild | 28.93 | 5.33 | 0.33 |
| 8 | 31-35 | Male | Mild | 35.06 | 3.41 | -1.45 |
| 9 | 51-55 | Male | Severe | 35.43 | 3.26 | -1.39 |
| 9 | 36-40 | Male | Mild | 36.67 | 2.45 | -2.14 |
| 10 | 71-75 | Male | Severe | 28.86 | 5.32 | 0.72 |
| 10 | 46-50 | Female | Mild | 28.78 | 5.48 | 0.87 |
| 11 | 26-30 | Female | Mild | 36.92 | 2.54 | -1.65 |
| 12 | 26-30 | Male | Moderate | 27.97 | 5.61 | 1.4 |
| 12 | 61-65 | Male | Severe | 32.49 | 4.29 | 0.18 |
| 13 | 66-70 | Female | Moderate | 25.3 | 6.36 | 2.3 |

### Conversion of viral load measurements to z-scores using viral load regression model

We recently developed a regression model fitted to viral load measurements within the first 15 days of symptoms across 16 datasets, capturing the viral load variation during the course of infection between different individuals (3). Here, to determine whether individual subjects in the current study had higher or lower than average viral load measurements relative to their duration of illness, we used the previously published regression model to calculate a z-score for each data point representing the deviation of the data point from the mean viral load trajectory i.e. the regression line (**Tables 1 and 2, Figures 1D and 1E**; see Methods). Viral load z-scores calculated from the data were not associated with the severity of illness (**Figure 1F)**. In our previous large-scale analysis of COVID-19 subjects (3), we showed that age and sex did not significantly influence URT viral load dynamics and that URT viral load dynamics did not affect the severity of illness. Therefore, in the present study, we did not adjust the viral load z-scores for these variables.

### Exploring molecular correlates of SARS-CoV-2 viral load using whole blood transcriptomics

We aimed to identify groups of genes for which expression correlated with viral load z-score, providing insights into the mechanisms controlling viral load. We first performed a gene signature-based deconvolution (13), as in our previous studies (11, 14). Interestingly, the computed proportion estimate of natural killer (NK) cell population was negatively correlated with viral load z-score (r = -0.62 and *P* = 0.010, **Supplementary Figure 1**). There was insufficient evidence to conclude a significant linear relationship between other leukocyte populations (B-cells, monocytes, neutrophils, CD4^+^ T-cells, and CD8^+^ T-cells) and viral load z-score. Gene expression counts were then adjusted for leukocyte mixture to remove the confounding effect of differences in blood leukocyte proportions between individuals. To make best use of the relatively small sample size of the selected 16 samples, we performed dimensionality reduction using weighted correlation network analysis (WGCNA) (15). First, we clustered the RNA-Seq profiles (n = 96) and removed an outlier (**Supplementary Figure 2**). Then, a gene co-expression network was constructed, and modules (clusters of highly co-expressed genes) were detected using the remaining 95 whole blood RNA-Seq profiles. Next, we correlated the first principal component of each module (module eigengene) to viral load z-scores in the group of 16 samples with paired data, reasoning that inducible mechanisms which restrict viral load would be enriched amongst the most strongly correlated modules. Twenty-four modules were significantly correlated with viral load (*P* < 0.01; **Figure 2, Table 3,** and **Supplementary Table 2**). To aid interpretation, we represented each module by its hub gene (gene with the highest connectivity within the module). Fourteen modules were positively correlated with viral load z-score and 10 were negatively correlated (**Figure 2A)**. *IFNA14* (Interferon Alpha-14) and *AIPL1* (Aryl Hydrocarbon Receptor Interacting Protein Like 1) modules showed the strongest negative correlation with viral load (r = -0.60 with *P* = 1e-10 and r = -0.60 with *P* = 2e-10, respectively). The largest positive correlation was observed for the *AC011455.2* module (r = 0.60, *P* = 2e-10).

**Figure 2.**
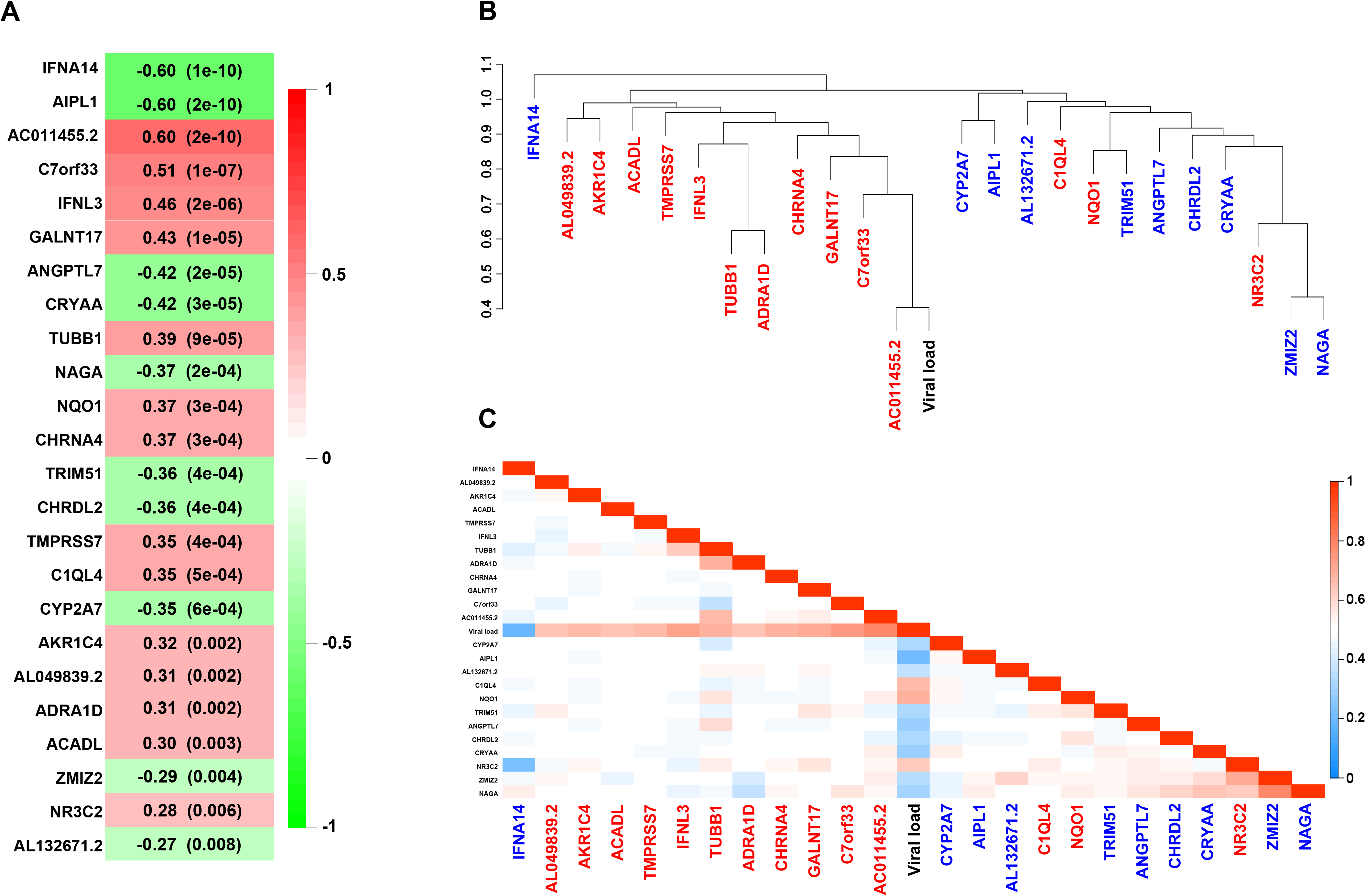
Peripheral blood gene expression modules correlated with viral load z-score. A) For each module, the Pearson correlation with viral load z-score and corresponding p-value are displayed. The Pearson correlation scale is depicted on the right. B and C) Module network and relationship with viral load z-score. The hierarchical clustering dendrogram of the module eigengenes (B) was generated using all genes in the modules and shows the dissimilarity of eigengenes with the distance measure being one minus correlation. Modules coloured in red and blue are, respectively, positively and negatively correlated with viral load. The heatmap (C) represents module eigengene adjacency calculated as (1 + correlation)/2.

**Table 3:**
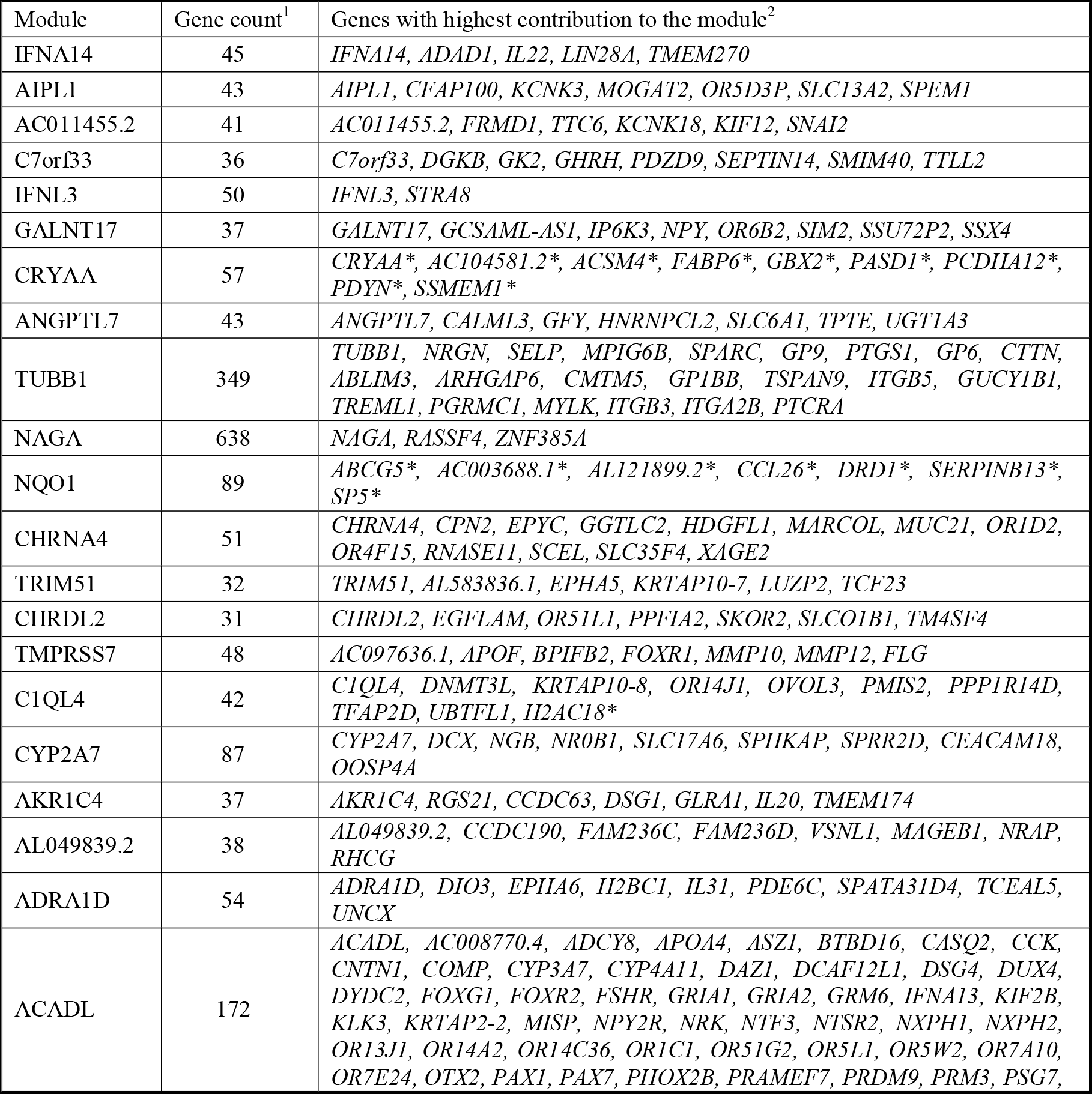
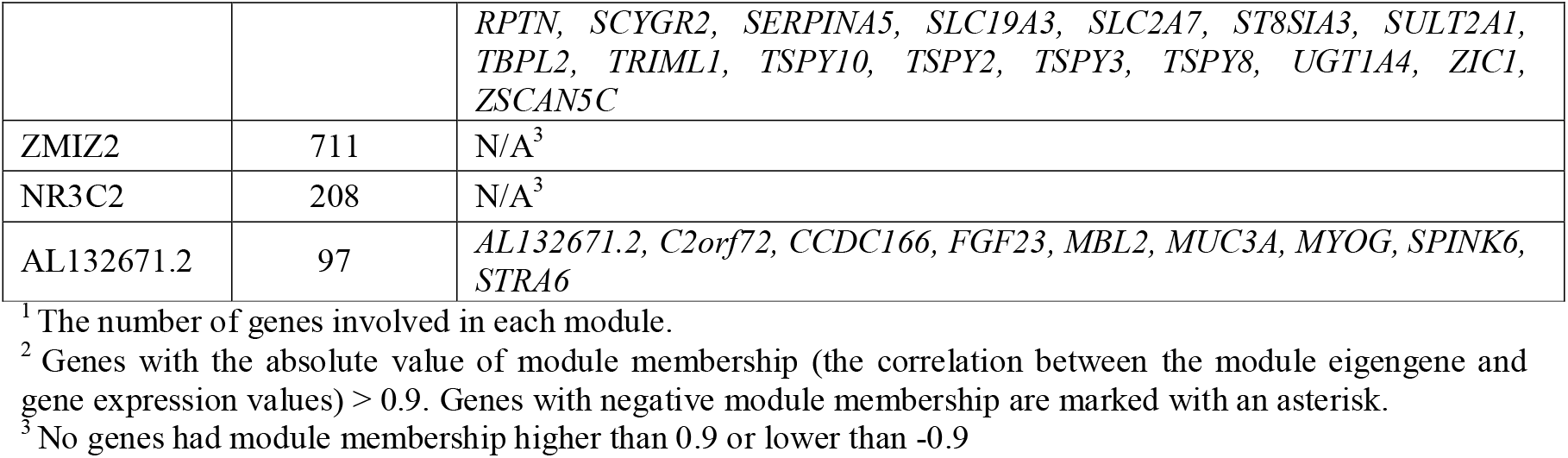
Modules significantly correlated with viral load z-score. Each module is represented by its hub gene. Complete lists of module genes and their information are provided in **Supplementary** Table 2.

We selected the top 6 significant modules for further analysis: *IFNA14*, *AIPL1*, *AC011455.2*, *C7orf33* (Chromosome 7 Open Reading Frame 33), *IFNL3* (Interferon Lambda 3), and *GALNT17* (Polypeptide N-Acetylgalactosaminyltransferase 17). *AC011455.2*, *C7orf33*, and *GALNT17* modules were positively correlated with viral load z-score and positioned very close to each other in the hierarchical clustering (**Figure 2B**). Therefore, we merged their gene sets to form a metamodule (*AC011455.2*/*C7orf33*/*GALNT17*; total gene count = 114) for further data analysis, assuming that the higher gene count would increase power to detect biologically relevant changes. We used Qiagen’s Ingenuity Pathway Analysis (IPA) for biological understanding of the modules (16). Correlation coefficients between module genes and viral load z-score were used to infer the activity pattern (activation or inhibition) of the biological processes involved such as enriched pathways and upstream regulators.

**Figure 3A** illustrates top enriched canonical pathways for the *IFNA14*, *AIPL1, AC011455.2/C7orf33/GALNT17,* and *IFNL3* modules (details provided in **Supplementary Table 3)**. Of these, 3 pathways showed an enrichment *P*-value of below 0.001 including ‘pathogen induced cytokine storm signalling’ (*P* = 5e-4) and ‘IL-22 (Interleukin-22) signalling’ (*P* = 8e-4) enriched in the *IFNA14* module, and ‘melatonin degradation’ (*P* = 2e-4) enriched in the *IFNL3* module. The pathogen induced cytokine storm signalling pathway encompasses the highest number of genes from the tested module (*IFNA14*, *IL22*, *CCL4* (C-C Motif Chemokine Ligand 4), *CD70*, and *COL4A4* (Collagen Type IV Alpha 4 Chain) from the IFNA14 module) compared to the other enriched pathways identified. *IFNA14* and *IL22*, two main members of the *IFNA14* module (module membership = 0.98 and *P* = 5e-64 for both genes), are key components of the pathway. Interestingly in our dataset both genes were negatively correlated with viral load z-score (r = -0.59 and *P* = 3e-10 for both genes), whereas *CCL4*, *CD70*, and *COL4A4* were positively correlated with viral load z-score. The IL-22 signalling pathway involves two genes, *IL22* and *IL22RA2* (Interleukin 22 Receptor Subunit Alpha 2), with a high contribution to the *IFNA14* module (module membership = 0.98 and 0.75 with *P* = 5e-64 and 1e-18, respectively) both negatively correlated with viral load z-score (r = -0.59 and -0.63 with *P* = 3e-10 and 7e-12, respectively). The superpathway of melatonin degradation includes three genes (*CYP2F1* (Cytochrome P450 Family 2 Subfamily F Member 1), *IL4I1* (Interleukin 4-induced gene-1), and *UGT1A1* (UDP Glucuronosyltransferase Family 1 Member A1) from the IFNL3 module. However, this pathway is likely to be of less importance here as the three genes each show relatively weak individual correlations with viral load z-score (**Supplementary Table 2**).

**Figure 3.**
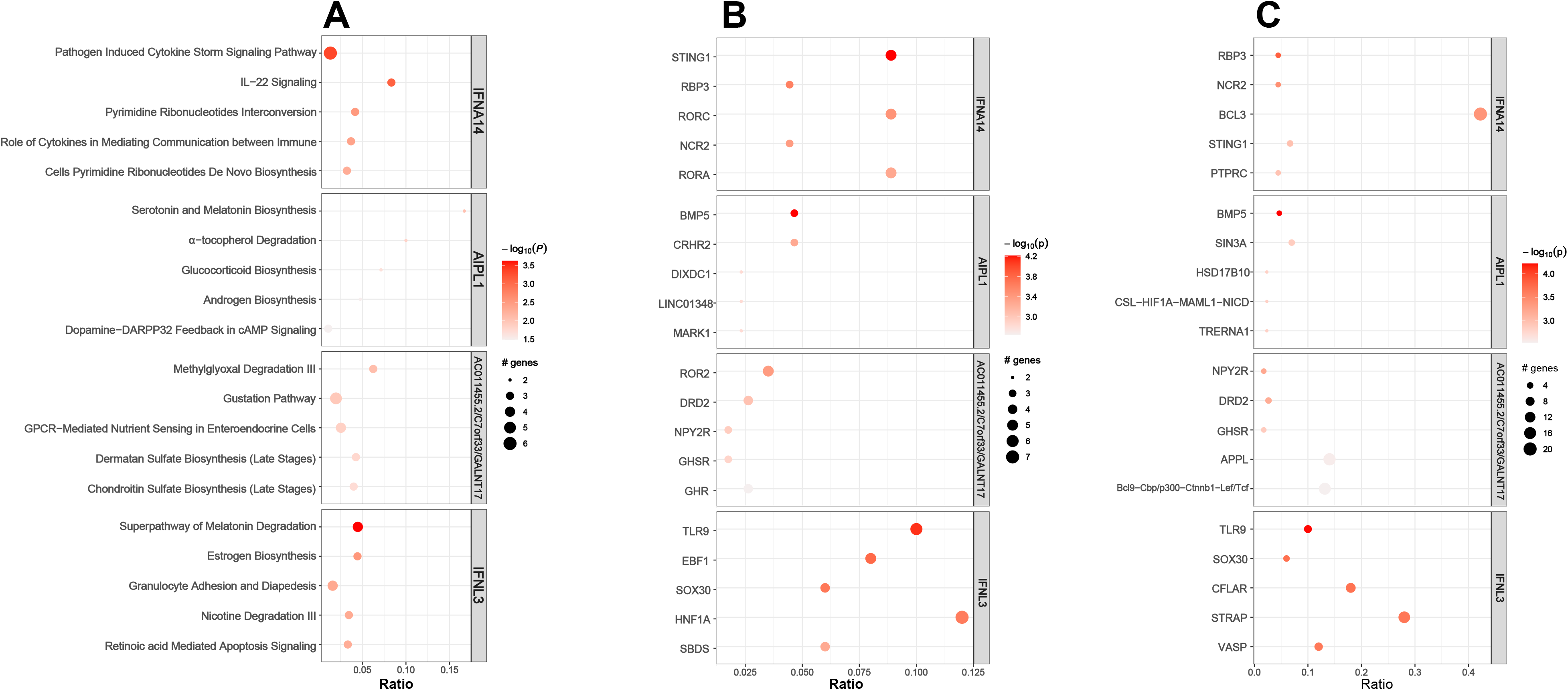
Ingenuity pathway analyses of peripheral blood gene expression modules most strongly correlated with viral load. A) For each module, the top 5 significant pathways are illustrated in descending order of statistical significance as indicated by colour. For each pathway, the size of the corresponding circle represents the number of module genes that map to the pathway. The x-axis shows the ratio of the number of genes common between the corresponding module and pathway divided by the total number of genes that map to the same pathway. B) and C) For each module, the 5 most significant upstream (B) and master regulators (C) are illustrated in descending order of statistical significance as indicated by colour. For each regulator, the size of the corresponding circle represents the number of module genes downstream to the regulator. The x-axis shows the ratio of the number of module genes downstream to the corresponding regulator divided by the total number of module genes.

*STING1* (Stimulator of Interferon Response CGAMP Interactor 1), *RBP3* (Retinol Binding Protein 3), and *RORC* (RAR Related Orphan Receptor C) were predicted to be the most significant upstream regulators of *IFNA14* module genes (*P* = 6e-5, 3e-4, and 4e-4, respectively) (**Figures 3B and 3C**). They interact with *IL22*, *IL22RA2*, *CCL4*, *CD70*, and *FEZ1* (Fasciculation and Elongation Protein Zeta 1), members of the *IFNA14* module which are mapped to the pathogen induced cytokine storm signalling and IL-22 signalling pathways (**Supplementary Tables 4 and 5**). None of the identified pathways and regulators showed reliable evidence of activation (absolute value of IPA activation z-score > 2) and therefore we were not able to infer an overall directionality (activation or inhibition) with respect to viral load.

We also applied the *BloodGen3Module* tool (17) to identify gene modules associated with viral load. Unlike WGCNA which detects modules from the analysed gene expression dataset, *BloodGen3Module* uses fixed functionally pre-annotated modules characterising different biological responses of distinct blood cell types. We used RNA-Seq data without adjustment for leukocyte-mixture and evaluated differential expression of these modules between samples with positive and negative viral load z-scores (n = 5 and n = 11, respectively). We identified an aggregate of five modules showing high ‘module response’ and higher module expression in subjects with positive viral load z-score (aggregate module A34; **Figure 4A and Supplementary Tables 6**). A module response is defined as the percentage of genes for a given module showing significant differential expression between the groups. From the module aggregate, the Prostanoids module showed the highest module response (97%). Interestingly, we observed a significant overlap between the A34 aggregate module and *TUBB1* (Tubulin Beta 1 Class VI) module which was found to be significantly positively correlated with viral load z-score by WGCNA (r = 0.39 with *P* = 9e-05; **Figure 2**). Seventy genes including *TUBB1* were common between the A34 and *TUBB1* modules (**Figure 4B and Supplementary Tables 6**) while A34 did not overlap with any other WGCNA module correlated with viral load z-score. Among the A34 modules, the Prostanoids module showed the highest overlap with the *TUBB1* module (from 36 genes involved in the Prostanoids module, 30 were also included in the *TUBB1* module).

**Figure 4.**
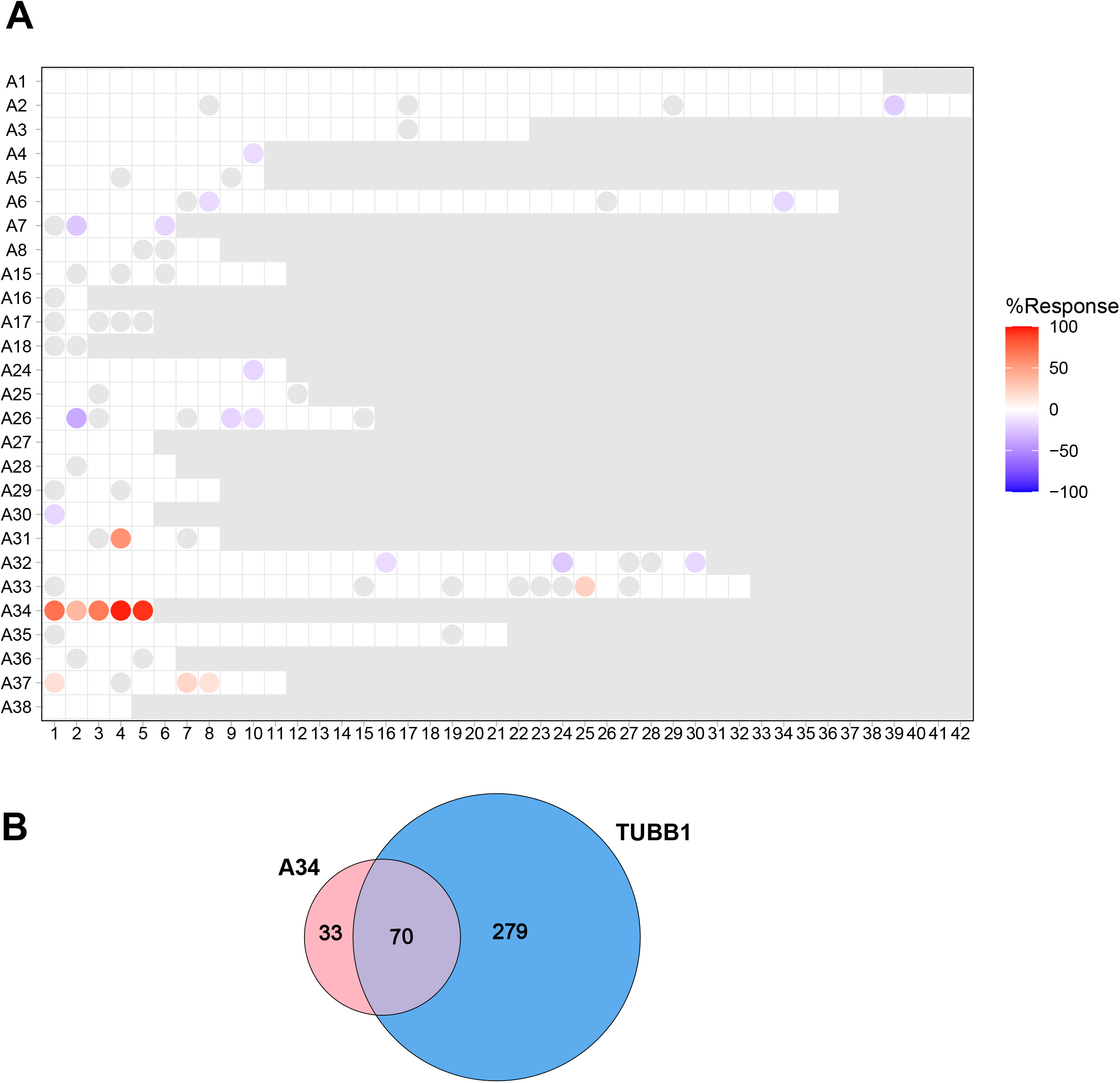
Pre-annotated blood gene expression modules associated with viral load. A) Module fingerprint grid plot. The differential expression of the modules is compared between two groups with positive and negative viral load z-score using t-test with fold change and p-value cut-off of 0.5 and 0.05, respectively. Each block corresponds to a module position. Each row represents a ‘module aggregate’ including modules with the same pattern of differential expression across reference datasets. Red and blue spots represent modules with increased and decreased abundance in the positive vs negative viral load z-score group, respectively. The gradient represents ‘module response’ which is the percentage of genes for a given module showing significant change in abundance between the two groups. Only modules with at least 15% response have been shown. B) Overlap of genes between A34 and *TUBB1* module.

### Exploring molecular correlates of SARS-CoV-2 viral load using NanoString assay of nasal epithelium

We analysed RNA isolated from nasal epithelium samples of 24 COVID-19 patients using a NanoString panel of 579 genes involved in core pathways and processes of human immune responses (**Supplementary Table 7**). Seventeen subjects also had paired URT viral load measurement within 15 days from symptom onset and had no evidence of bacterial co-infection. Using WGCNA we identified seven gene co-expression networks which we refer to as “pseudo-modules” since they were detected using a relatively low number of genes included in the NanoString panel (**Supplementary Table 8)**. Only one pseudo-module was correlated with viral load z-score at significance threshold of 0.05 (*PTK2* (Protein Tyrosine Kinase 2) module; *P* = 0.016). Additionally, correlation analysis between individual gene expression and viral load z-score detected significant correlation (absolute correlation coefficient > 0.5 and *P* < 0.05; **Figure 5**) in 12 individual genes, 11 of which were positively correlated and one (*GNLY* (Granulysin)) was negatively correlated.

**Figure 5.**
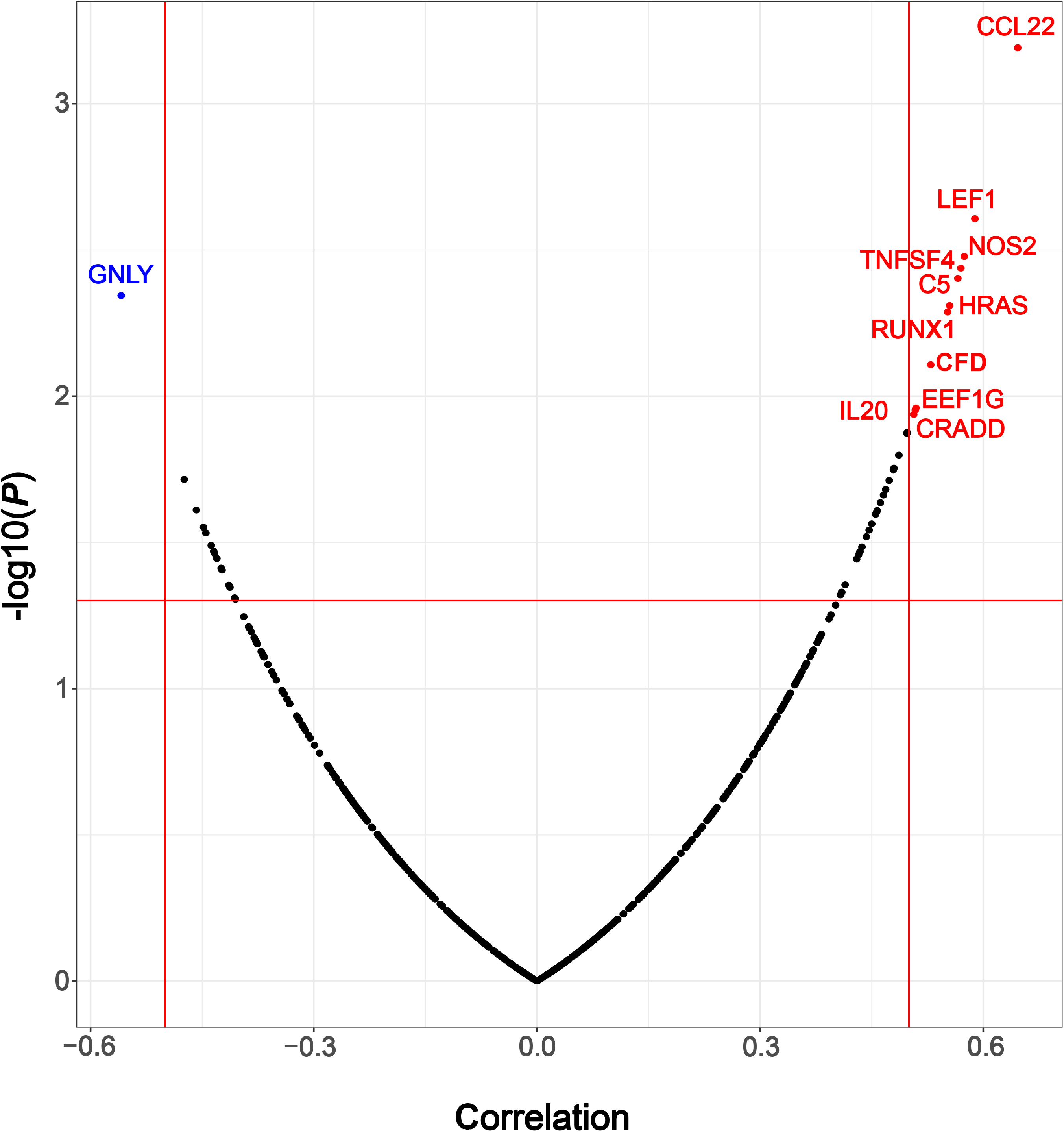
Correlation between nasal epithelium transcriptome and viral load z-score. The volcano plot illustrates correlation coefficients and corresponding p-values. Each dot represents a gene included in the NanoString panel. Genes strongly correlated with viral load z-score (absolute correlation coefficient > 0.5 and *P* < 0.05) are coloured as red (positive correlation) and blue (negative correlation).

## Discussion

Understanding mechanisms controlling SARS-CoV-2 viral load in the URT can provide valuable leads towards treatment and vaccine strategies aimed at reducing viral transmission. Such strategies have recently been highlighted as potential “game changers” as societies adapt to living with COVID-19 (18). Current evidence suggests that the mechanisms controlling URT viral load may be different from those controlling LRT viral load and disease severity (3–5). However, our current knowledge concerning control of URT viral load is far from complete. To unravel the biological complexity underlying the control of SARS-CoV-2 viral load, we sought to identify correlates of the variation in viral load which occurs in naturally infected individuals. Of note, we used samples from the first wave of infection in Europe, prior to vaccination, infection-induced immunity, and circulation of important variants of SARS-CoV-2. To account for the dynamic nature of URT viral load, which rapidly increases to a peak just before symptom onset and then declines more slowly, we quantified viral loads by their standardised deviation (z-scores) from a previously-derived average trajectory (3). We correlated viral load z-scores with paired peripheral blood and nasal epithelium transcriptomes.

After excluding individuals with proven or suspected bacterial co-infection and URT SARS-CoV-2 viral load samples taken more than 15 days after symptom onset, relatively small numbers of subjects for blood and nasal transcriptome analysis (n = 16 and n = 17, respectively) remained. This prompted the use of gene modules rather than individual genes for our primary analysis. This would reduce the complexity of large gene networks into relevant modules and increase the statistical power to detect those correlating with viral load. An individual module compromises genes that are more densely connected than expected by chance and often involved in the same biological functions (19). We applied two different methods to detect gene clusters, *WGCNA* and *BloodGen3Module*. The first identifies modules directly from the gene expression data and the later uses pre-annotated modules.

The peripheral blood module most significantly associated with URT viral load, had *IFNA14* as its hub gene. *IFNA14* encodes the type I interferon, interferon α14. Interferons are glycoprotein cytokines made and released by host lymphocytes and considered to be key effectors in antiviral responses. However, their pattern of expression and function during SARS-CoV-2 infection is controversial. While some studies suggest protective effects of interferons in severe COVID-19 (20–22), others indicate poor clinical outcomes in those with increased production of interferons (23–26). There is limited data available on the correlation between SARS-CoV-2 viral load and interferon expression. Sposito et al. evaluated nasopharyngeal swabs of COVID-19 patients and showed that the expression of type I and III interferons was significantly associated with viral load in patients under 70 years old (26). However, those aged over 70 years showed no association and/or showed a significantly lower correlation coefficient. This evaluation did not include *IFNA14*. Also, it appears that their viral load measurements were not adjusted for the time between sample collection and symptom onset. *IFNA14* has been shown to activate a potent antiviral response *via* binding to IFNAR1 and IFNAR2 (Interferon Alpha and Beta Receptor Subunits 1 and 2) receptors (27, 28). This triggers the activation of JAK/STAT (Janus Kinase/Signal Transducer and Activator of Transcription) signalling complexes which subsequently induces the expression of ISGs (interferon-stimulated genes) that inhibit virus infection (29). The strong negative correlation between the *IFNA14* module as well as *IFNA14* as an individual gene and viral load in our data suggests that IFNA14 signalling could play a key role in controlling SARS-CoV-2 viral load, i.e. increased expression of *IFNA14* restricts viral replication. Schuhenn et al. recently showed that IFNA14 is one of the most potent interferon alpha subtypes inhibiting SARS-CoV-2 replication and can cause a significant reduction of SARS-CoV-2 viral titre by up to 10^5^-fold (30). Furthermore, unpublished data suggest that, compared to IFNA2 (Interferon Alpha-2), which was used to treat COVID-19 patients in an uncontrolled exploratory study in China (31), IFNA14 is more efficient at preventing the infection while less detrimental to the immune system (32). Not only is *IFNA14* important as an individual gene, but it also represents a network of highly connected genes in our data, the *IFNA14* module, which showed a high enrichment of two canonical pathways ‘pathogen induced cytokine storm signalling’ and ‘IL-22 signalling’. Although SARS-CoV-2 can trigger a ‘cytokine storm’ (33, 34), the changes in expression of genes in this pathway were not consistently associated with activation or inhibition of the pathway, agreeing with previous findings that ability to control of URT viral load is dissociated from severity of illness (3, 4). IL-22 is a cytokine released by several immune cells such as Th22 (T helper cells type 22) and plays an important role at mucosal barriers, orchestrating the interaction between the epithelial cell layer and local immune system in response to infections (35). IL-22 stimulates the IL-22 receptor complex on epithelial cells resulting in downstream activation of JAK-STAT signalling pathway which induces multiple antiviral responses and therefore can be protective during SARS-CoV-2 infection (35–37). Elevated levels of IL-22 in the plasma have been implicated as a hallmark of severe COVID-19 (24). Taken together there is compelling evidence that the genes in the *IFNA14* module act to reduce URT viral load, and add to the evidence that interferon α14 should be considered as a candidate treatment to reduce viral load in the URT and decrease transmissibility of SARS-CoV-2.

The *AIPL1* module was the second top module negatively corelated with viral load z-score. Unlike *IFNA14*, *AIPL1* is not known to be involved with the pathogenesis of COVID-19, and the enriched pathways for this module contained relatively few genes. Nevertheless, enrichment of the ‘α-tocopherol degradation’ pathway suggests a potential role of α-tocopherol (also known as Vitamin E) in the control of viral load. α-tocopherol is an antioxidant which may enhance the function of innate and adaptive immune cells, for example increasing NK cell activity and the phagocytic capacity of leukocytes, which could bolster the immune response to reduce pathogen load as observed in influenza (38, 39). Emerging evidence suggests that water soluble derivatives of α-tocopherol have a potent antiviral response especially when they are used synergistically with remdesivir to inhibit SARS-CoV-2 RNA-dependent RNA polymerase (40).

We also identified modules positively correlated with viral load, possibly indicating that these modules are induced in response to increasing amounts of virus or that expression of these genes favour an increase in viral replication. The most significant of these modules was the *IFNL3* module. The hub gene, *IFNL3*, encodes a type III interferon which is a cytokine activated in response to mucosal viral infections and signals through the heterodimeric IFNLR (Interferon Lambda Receptor) that is expressed distinctly in the URT epithelial cells. This stimulates the activation of several transcription factors which upregulate ISGs. Type III interferon signalling pathway is considered slower and induces a weaker ISG response than type 1 interferons (41, 42). The most significant upstream regulator of the *IFNL3* module is TLR9 (Toll Like Receptor 9), which may be stimulated by unmethylated CpG (Cytosine-phosphate-Guanine) sequences during SARS-CoV-2 infection (43) and result in the observed upregulation of *IFNL3* module genes.

Using *BloodGen3Module*, we identified a cluster of blood pre-annotated transcriptional modules (A34) positively correlated with viral load. This was particularly interesting as these modules showed a significant overlap with the *TUBB1* module found to be significantly positively correlated with viral load by WGCNA. From the A34 modules, the Prostanoids module showed the highest module response and overlap with the *TUBB1* module. Prostanoids are a subclass of eicosanoids and regulate the inflammatory response (44). The observed association between the prostanoids module expression and viral load z-score suggests that high levels of prostanoids may supress processes which constrain viral load and therefore promote high viral load levels. This is supported by a recent study showing that abrogation of eicosanoid signalling reduces viral load and rescues mice from fatal SARS-CoV-2 infection (45).

In addition to peripheral blood samples, we studied samples taken from the primary infection site, the nasal epithelium. Both blood and nasal transcriptomes can reflect the host immune response to the infection. In a respiratory infection, epithelial cells are directly infected, and peripheral blood leukocytes also respond to signals arising from the site of infection (46). However, the difference in the transcriptomic analysis approach we used for each dataset (RNAseq for peripheral blood and NanoString assay for nasal samples) made it difficult to compare the results directly. The NanoString assay analysed a relatively small number of genes (579 genes involved in immune response) and therefore the data did not yield reliable module level results. However, individual genes correlated with URT viral load z-score were identified that may be of interest. *GNLY* was the only negatively correlated gene representing a likely role in the control of viral load. It is produced by a variety of killer cells such as cytotoxic T lymphocytes and NK cells, and it has both cytolytic and proinflammatory activity (47). Indeed, the expression of *GNLY* in lymphocytes has been reported to be associated with recovery in COVID-19 suggesting it may play a major role in clearance of infected cells and termination of infection (48). In agreement with the correlation between *GNLY* and viral load, we also showed that cell proportion estimates of NK cells in peripheral blood were negatively correlated with viral load z-score, highlighting the importance of this cell population in constraining the virus. For example, Witkowski et al. showed COVID-19 patients with normal NK cell numbers demonstrated a more rapid decline of viral load compared to those with low NK cell numbers (49).

Our study was limited by the relatively small sample size which may have reduced the statistical power and resulted in missed opportunities to capture some biological signals. Additionally, we cannot establish from this data whether the molecular mechanisms identified are cause or consequence of the viral load, although there are plausible mechanisms which suggest causal roles in some cases.

## Conclusions

To our knowledge, this is the most comprehensive study focusing on identifying molecular correlates of the SARS-CoV-2 viral load control in the URT. We identified numerous molecular processes which may contribute to the control of URT viral load. These candidate mechanisms can be the focus of further functional studies and may lead to new strategies to prevent COVID-19 and reduce SARS-CoV-2 transmission.

## Methods

### Study design and participants

We studied 82 COVID-19 patients recruited through the GEN-COVID study (www.gencovid.eu), a multi-center and prospective cohort designed to evaluate the effect of genetic factors on SARS-CoV-2 infection. Subjects were recruited at Hospital Clínico Universitario de Santiago de Compostela (Galicia, Spain) between March 2020 and May 2020, during the first wave of infections in Spain, before significant levels of infection- and vaccine-induced immunity in the community. COVID-19 was defined according to the Spanish national guidelines (https://www.mscbs.gob.es/profesionales/saludPublica/ccayes/alertasActual/nCov/documentos.htm). The severity of the disease was defined as mild, moderate, and severe based on WHO scoring for COVID-19 patients and as described previously (50, 51). We also included 18 uninfected controls, and 9 subjects with non-COVID-19 infections recruited through the PERFORM Consortium.

### Sample collection

Blood samples and nasal epithelium specimens were collected at the same time at hospital for moderate and severe COVID-19 subjects and at home for subjects with mild disease. Whole blood was collected into PAXgene blood RNA tubes (PreAnalytiX) and nasal epithelium samples were collected in Oragene CP-190 kit (DNA Genotek). Samples were processed as described previously (51, 52).

One COVID-19 subject contributed two paired sets of samples (viral load and blood RNA-Seq; **Table 1**) collected 3 days apart. We included both as they showed a noticeable difference in viral load z-score and hence were informative.

### RNA isolation

Total RNA was isolated from blood and nasal epithelium samples using PAXgene blood miRNA extraction and RNeasy microkit, respectively, according to the manufacturer’s protocols (Qiagen). RNA amount and integrity were assessed using TapeStation 4200 (Agilent). RNA quality was checked based on DV200 metric to ensure that sufficient percentage (over 50%) of RNA fragments were greater than 200 nucleotides in length and also to estimate the optimal sample input for the *nCounter* NanoString analysis.

### Viral load measurements

#### Viral load quantification

Nasopharyngeal samples were collected in Universal Transport Medium (UTM) tubes and assessed for the presence and viral load of SARS-CoV-2. We detected viral particles using a multiplex real-time PCR with the Allplex™SARS-CoV-2 Assay (Seegene). Viral load values (viral copies per ml) were computed from the Ct values as described previously (3).

#### Calculation of viral load z-scores

A regression model of the average trajectory of viral load over time and quantification of variation between individuals, using data from 16 datasets, was reported previously (3). Viral load values from the present study were compared to the regression line to assess whether a particular viral load measurement, sampled a certain number of days after symptom onset, was higher or lower than average. A ‘z-score’ was calculated for each data point by calculating its deviation from the mean trajectory and dividing by the standard deviation of the variation in viral load around the mean trajectory (**Fig. 1D**).

### RNA sequencing

Paired-end sequencing was performed at The Wellcome Centre for Human Genetics in Oxford, UK as described previously (51). Sequencing was carried out using Novaseq6000 platform providing 150 bp paired end reads.

#### RNA-Seq upstream analyses

Adapter trimming and quality control of sequencing reads were performed with Trimmomatic version 0.36 and FastQC version 0.11.7, respectively (53, 54). The reads were then mapped against hg38 reference genome using STAR version 2.7.1a (55). RSEM version 1.3.1 was used for transcript quantification (56). Next, we performed a gene signature-based deconvolution using CellCODE as in our previous work and adjusted gene expression for leukocyte (B cells, monocytes, neutrophils, NK cells, CD4^+^ T cells, and CD8^+^ T cells) mixture (11, 13, 14).

### NanoString experiment

#### NanoString *n*Counter assay

We analysed immunological gene expression profiles of nasal epithelium using the *SPRINT nCounter* system (NanoString Technologies) with the Human Immunology V2 Panel (579 genes covering the core pathways and processes of the immune response, and 15 internal reference genes for data normalization). The detail of the assay is described previously (52).

#### Differential gene expression analysis

The gene expression counts adjusted for leukocyte mixture were correlated with viral load z-scores using edgeR (57).

#### Weighted correlation network analysis

Gene counts were normalised using variance stabilizing transformation (*VST*) function of DESeq2 R package (58) and adjusted for leukocyte mixture using *removeBatchEffect* function of limma R package (59). We used WGCNA version 1.71 R package for weighted correlation network analysis (15).

### Module repertoire analysis

We applied *BloodGen3Module* version 1.4.0 R package (17) to the normalised gene expression counts unadjusted for cell-mixture from 16 samples with paired viral load, collected in the absence of bacterial co-infection. The package encompasses 382 functionally annotated blood transcriptional modules which have been grouped into 38 “aggregates” (A1-A38). The differential expression of the modules was compared between two groups with positive and negative viral load z-scores (n = 5 and n = 11, respectively) using t-test with fold change and p-value cut-off of 0.5 and 0.05, respectively. For each module, we computed ‘module response’ as the percentage of genes for the module showing significant differential expression between the two groups.

### Further statistical analysis

The normality of distributions was assessed using the Shapiro-Wilk normality test. Pearson correlation was used to analyse the degree of association between two continuous variables. An independent-samples t-test and one-way ANOVA with Tukey’s post hoc test were used to compare continuous variables between two and multiple groups, respectively.

## Data Availability

Raw RNA-Seq data and corresponding metadata are available on ArrayExpress under the accession E-MTAB-12791. NanoString nCounter data and corresponding metadata are available at https://github.com/MahdiMoradiMarjaneh/COVID19_viral_load.

## List of abbreviations

AIPL1: Aryl Hydrocarbon Receptor Interacting Protein Like 1
C7orf33: Chromosome 7 Open Reading Frame 33
CCL4: C-C Motif Chemokine Ligand 4
COL4A4: Collagen Type IV Alpha 4 Chain
COVID-19: coronavirus disease
CpG: Cytosine-phosphate-Guanine
Ct value: cycle threshold value
CYP2F1: Cytochrome P450 Family 2 Subfamily F Member 1
FEZ1: Fasciculation and Elongation Protein Zeta 1
GALNT17: Polypeptide N-Acetylgalactosaminyltransferase 17
GNLY: Granulysin
IFNA14: Interferon Alpha 14
IFNA2: Interferon Alpha-2
IFNAR1 and IFNAR2: Interferon Alpha and Beta Receptor Subunits 1 and 2
IFNL3: Interferon Lambda 3
IFNLR: Interferon Lambda Receptor
IL-22: Interleukin-22
IL22RA2: Interleukin 22 Receptor Subunit Alpha 2
IL4I1: Interleukin 4-induced gene-1
IPA: Ingenuity Pathway Analysis
ISGs: interferon-stimulated genes
JAK/STAT: Janus Kinase/Signal Transducer and Activator of Transcription
LRT: lower respiratory tract
NK cells: natural killer cells
PCA: principal component analysis
PTK2: Protein Tyrosine Kinase 2
RBP3: Retinol Binding Protein 3
RNA-Seq: RNA sequencing
RORC: RAR Related Orphan Receptor C
SARS-CoV-2: Severe Acute Respiratory Syndrome Coronavirus 2
STING1: Stimulator of Interferon Response CGAMP Interactor 1
Th22: T helper cells type 22
TLR9: Toll Like Receptor 9
TUBB1: Tubulin Beta 1 Class VI
UGT1A1: UDP Glucuronosyltransferase Family 1 Member A1
URT: upper respiratory tract
VL: viral load
WGCNA: weighted correlation network analysis

## Declarations

### Ethics approval and consent to participate

The GEN-COVID and PERFORM (Personalised Risk assessment in Febrile illness to Optimise Real-life Management across the European Union; perform2020.org/) studies were conducted according to the guidelines of the Declaration of Helsinki and approved by the Ethics Committee of Galicia (CEIC, ref 2020/178, 18/03/2020) and St Mary’s Research Ethics Committee (16/LO/1684, 25/02/2013), respectively. A written informed consent was obtained for each participant. If this was not done at the time of sampling, a retrospective consent was sought at the earliest appropriate opportunity.

Here, we have replaced sample and subject IDs with identifiers that cannot reveal the identity of the study subjects.

### Consent for publication

Not applicable

### Availability of data and materials

Raw RNA-Seq data and corresponding metadata are available on ArrayExpress under the accession E-MTAB-12791. NanoString nCounter data and corresponding metadata are available at https://github.com/MahdiMoradiMarjaneh/COVID19_viral_load. Codes used in the analyses can be accessed on the same GitHub repository.

### Competing interests

The authors declare that they have no competing interests.

### Funding

This work was supported by UKRI (MRC) and the DHSC (NIHR) (Grant Ref: MR/V027409/1). This study also received support from Instituto de Salud Carlos III ([ISCIII] TRINEO: PI22/00162; DIAVIR: DTS19/00049; Resvi-Omics: PI19/01039 [AS]; ReSVinext: PI16/01569 [F.M.-T.]; Enterogen: PI19/01090 [F.M.-T.]; OMI-COVI-VAC (PI22/00406 [F.M.-T.] cofinanciados FEDER), GAIN: Grupos con Potencial de Crecimiento (IN607B 2020/08, [A.S.]); ACIS: BI-BACVIR (PRIS-3, [A.S.]), and CovidPhy (SA 304 C, [A.S.]); and consorcio Centro de Investigación Biomédica en Red de Enfermedades Respiratorias (CB21/06/00103; F.M.-T.); GEN-COVID (IN845D 2020/23, F.M.-T.) and Grupos de Referencia Competitiva (IIN607A2021/05, F.M.-T.). The funders were not involved in the study design, collection, analysis, interpretation of data, the writing of this article or the decision to submit it for publication.

M.M.M. is supported in part by the NIHR Biomedical Research Centre of Imperial College NHS Trust.

JD.C. and L.C.O. acknowledge funding from the MRC Centre for Global Infectious Disease Analysis (reference MR/R015600/1), jointly funded by the UK Medical Research Council (MRC) and the UK Foreign, Commonwealth & Development Office (FCDO), under the MRC/FCDO Concordat agreement and is also part of the EDCTP2 programme supported by the European Union.

F.L. is supported by an MRC clinical training fellowship [award MR/W000970/1]. F.L. and R.T. are supported by the UK Coronavirus Immunology Consortium (UKCIC).

H.R.J. received support from the Wellcome Trust (4-year PhD programme, grant number 215214/Z/19/Z).

M.K. acknowledges support from the Wellcome Trust and the Medical Research Foundation Grants (206508/Z/17/Z and MRF-160-0008-ELP-KAFO-C0801).

L.C.O. declares grant funding from Merck Group on an unrelated project.

### Authors’ contributions

Conceptualization: A.J.C, J.D.C., L.C.O., R.S.T.; Data curation: M.M.M., I.R.-C., A.G.-C., D.H.-C., H.R.J., C.Y.F., Y.W., F.L.; Formal analysis: M.M.M., J.D.C, A.S., A.G.-C., M.K.; funding acquisition: A.J.C, J.D.C, L.C.O., R.S.T., M.L., M.K., F.M.-T., I.R.-C., A.G.-C., A.S.; Investigation: I.R.-C., A.G.-C., V.J.W., S.N., G.D’S., D.H.-C.; Methodology: M.M.M., J.D.C., A.G.-C., A.S., H.R.J., V.J.W., D.H.-C.; Project administration: A.J.C, A.S., A.G.-C., F.M.-T.; Resources: A.J.C., F.M.-T., I.R.-C., A.G.-C., M.K.; Software: M.M.M, H.R.J., D.H.-C.; Supervision: A.J.C, L.C.O., R.S.T., M.K., V.J.W., M.L., F.M.-T.; Validation: M.M.M.; Visualisation: M.M.M.; Writing—original draft: M.M.M., A.J.C..; Writing—review and editing: ALL.

## Acknowledgements

The members and affiliations of the PERFORM consortium and the GEN-COVID (www.gencovid.eu) study group are listed in the Supplementary text file.

## Notes

### Competing Interest Statement

The authors have declared no competing interest.

### Author Declarations

The GEN-COVID and PERFORM (Personalised Risk assessment in Febrile illness to Optimise Real-life Management across the European Union; perform2020.org/) studies were conducted according to the guidelines of the Declaration of Helsinki and approved by the Ethics Committee of Galicia (EL COMITE ETICO DE INVESTIGACION CLINICA (CEIC) Galicia, ref 2020/178, 18/03/2020) and St Mary's Research Ethics Committee (16/LO/1684, 25/02/2013), respectively.

