## Supplementary Figures for "Analysis of blood and nasal epithelial transcriptomes to identify mechanisms associated with control of SARS-CoV-2 viral load in the upper respiratory tract"

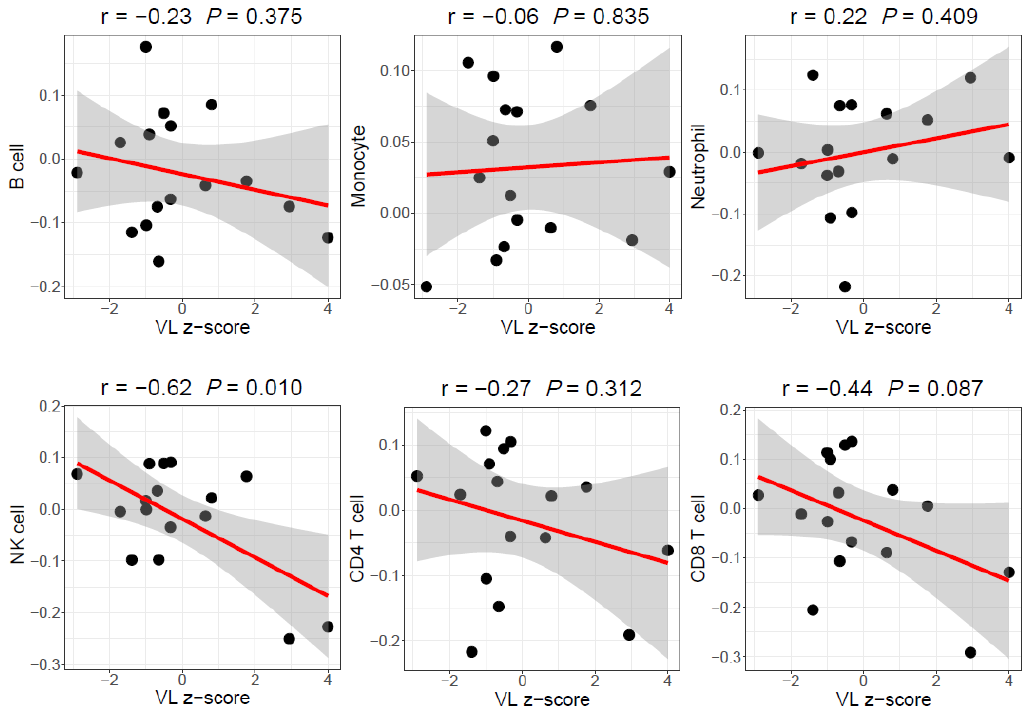

**Supplementary Figure 1**. Correlation between computed proportion estimates of blood leukocyte populations and URT viral load. For each population, proportion estimates are plotted against viral load z-scores. Each dot represents a sample with paired viral load and blood transcriptome data. The gray shading represents the 95% confidence interval around the line of best fit (red).

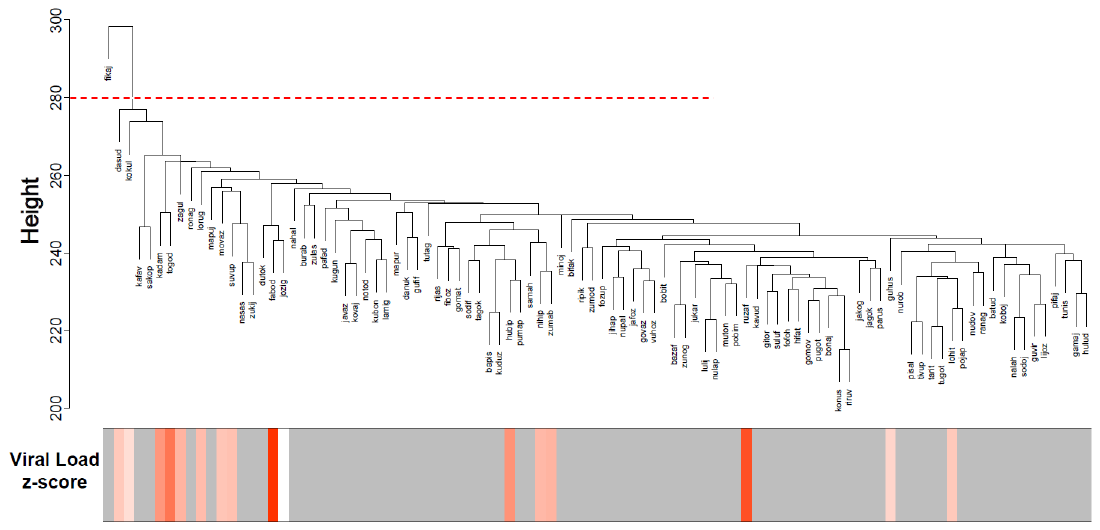

**Supplementary Figure 2**. Sample clustering and viral load z-scores. The dendrogram represents the clustering of RNA-Seq profiles. The red dashed line is the height cut-off to detect outliers. The lower panel shows the z-score heatmap with white being corresponded to a low value, red a high value, and grey a missing entry.
