## Supplementary Text for "Analysis of blood and nasal epithelial transcriptomes to identify mechanisms associated with control of SARS-CoV-2 viral load in the upper respiratory tract"

**Members of the GEN-COVID (www.gencovid.eu) study group**

Antonio Aguilera Guirao^15^, Julián Álvarez Escudero^16^, Antonio Antela López^17^, Gema Barbeito-Castiñeiras^9^, Xabier Bello Paderne^6^, Miriam Ben García^6^, María Victoria Carral García^18^, Miriam Cebey López^6^, Amparo Coira Nieto^15^, Mónica Conde Pájaro^19^, José Javier Costa-Alcalde^15^, María José Currás Tuala^6^, Ana Isabel Dacosta Urbieta^6,8^, Blanca Díaz Esteban^8^, María Jesús Domínguez Santalla^17^, Cristina Fernández Pérez^19^, Juan Fernández Villaverde^20^, Cristóbal Galbán Rodríguez^20^, José Luis García Allut^20^, Luisa García Vicente^6^, Elena Giráldez Vázquez^20^, Alberto Gómez-Carballa^5,6,7^, José Gómez Rial^6,8,14^, Francisco Javier González Barcala^21^, Beatriz Guerra Liñares^19^, Pilar Leboráns-Iglesias^8^, Beatriz Lence Massa^20^, Marta Lendoiro Fuentes^6,8^, Montserrat López Franco^6,8^, Ana López Lago^20^, Federico Martinón‑Torres^6,7,8^, Daniel Navarro de la Cruz^15^, Eloína Núñez Masid^22^, Juan Bautista Ortola Devesa^23^, Jacobo Pardo Seco^6^, María Pazo Núñez^17^, Marisa Pérez del Molino Bernal^15^, Hugo Pérez Freixo^19^, Lidia Piñeiro Rodríguez^6^, Sara Pischedda^6^, Manuel Portela Romero^5^, Antonio Pose Reino^18^, Gloria María Prada Hervella^16^, Teresa Queiro Verdes^19^, Lorenzo Redondo Collazo^6,8^, Patricia Regueiro Casuso^6^, Susana Rey García^6,8^, Sara Rey Vázquez^6^, Vanessa Riveiro Blanco^21^, Irene Rivero-Calle^6,7,8^, Carmen Rivero Velasco^20^, Nuria Rodríguez Núñez^21^, Carmen Rodríguez‑Tenreiro Sánchez^6^, Eva Saborido Paz^20^, José Miguel Sadiki Orayyou^6^, Carla Saito Villanueva ^20^, Sonia Serén Fernández^6^, Pablo Souto Sanmartín^19^, Manuel Taboada Muñiz^16^, Rocío Trastoy Pena^15^, Mercedes Treviño Castellano^15^, Luis Valdés Cuadrado^21^, Pablo Varela García^17^, María Soledad Vilas Iglesias^6^, Sandra Viz Lasheras^6^, Rocio Ferreiro‑Iglesias^24^, Iria Bastón‑Rey^24^ & Cristina Calviño‑Suárez^24^

^14^Laboratorio de Inmunologìa, Servicio de Análisis Clìnicos, Hospital Clìnico Universitario Santiago de Compostela, Servizo Galego de Saúde, Galicia, Spain

^15^Microbiology Department, Hospital Clìnico Universitario de Santiago de Compostela, Galicia, Spain

^16^Anesthesiology and Resuscitation Service, Hospital Clínico Universitario de Santiago, Santiago de Compostela, Galicia, Spain

^17^Internal Medicine Service, Hospital Clínico Universitario de Santiago, Santiago de Compostela, Galicia, Spain

^18^Director of Nursing Processes, Hospital Clínico Universitario de Santiago, Santiago de Compostela, Galicia, Spain

^19^Preventive Medicine Department, Hospital Clínico Universitario de Santiago de Compostela, Galicia, Spain

^20^Intensive Medicine Department, Hospital Clìnico Universitario de Santiago de Compostela, Galicia, Spain

^21^Pneumology Department, Hospital Clìnico Universitario de Santiago de Compostela, Galicia, Spain

^22^Manager of the Health Care Area of Santiago de Compostela and Barbanza, Hospital Clínico Universitario de Santiago, Santiago de Compostela, Galicia, Spain

^23^Clinical Chemistry Laboratory, Hospital Clínico Universitario de Santiago, Santiago de Compostela, Galicia, Spain

^24^Digestive Department, Hospital Clìnico Universitario de Santiago de Compostela, Galicia, Spain

**Members of the PERFORM Consortium**

**PARTNER: IMPERIAL COLLEGE (UK)**

Chief investigator/PERFORM coordinator:

Michael Levin

Principal and co-investigators; work package leads (alphabetical order)

Aubrey Cunnington (grant application)

Tisham De (work package lead)

Jethro Herberg (Principle Investigator, Deputy Coordinator, grant application)

Myrsini Kaforou (grant application, work package lead)

Victoria Wright (grant application, Scientific Coordinator)

Research Group (alphabetical order)

Lucas Baumard; Evangelos Bellos; Giselle D’Souza; Rachel Galassini; Dominic Habgood-Coote; Shea Hamilton; Clive Hoggart; Sara Hourmat; Heather Jackson; Ian Maconochie; Stephanie Menikou; Naomi Lin; Samuel Nichols; Ruud Nijman; Ivonne Pena Paz; Priyen Shah; Ching-Fen Shen; Clare Wilson

Clinical recruitment at Imperial College Healthcare NHS Trust (alphabetical order)

Amina Abdulla; Ladan Ali; Sarah Darnell; Rikke Jorgensen; Sobia Mustafa; Salina Persand

Imperial College Faculty of Engineering

Molly Stevens (co-investigator), Eunjung Kim (research group); Benjamin Pierce (research group)

Clinical recruitment at Brighton and Sussex University Hospitals

Katy Fidler (Principle Investigator)

Julia Dudley (Clinical Research Registrar)

Research nurses: Vivien Richmond, Emma Tavliavini

Clinical recruitment at National Cheng Kung University Hospital

Ching-Fen Shen (Principal Investigator); Ching-Chuan Liu (Co-investigator); Shih-Min Wang (Co-investigator), funded by the Center of Clinical Medicine Research, National Cheng Kung University

**PARTNER: SERGAS (Spain)**

Principal Investigators

Federico Martinón-Torres^1^

Antonio Salas^1,2^

Research Group (alphabetical order)

Fernando Álvez González^1^, Cristina Balo Farto^1^, Ruth Barral-Arca^1,2^, María Barreiro Castro^1^, Xabier Bello^1,2^, Mirian Ben García^1^, Sandra Carnota^1^, Miriam Cebey-López^1^, María José Curras-Tuala^1,2^, Carlos Durán Suárez^1^, Luisa García Vicente^1^, Alberto Gómez-Carballa^1,2^, Jose Gómez Rial^1^, Pilar Leboráns Iglesias^1^, Federico Martinón-Torres^1^, Nazareth Martinón-Torres^1^, José María Martinón Sánchez^1^, Belén Mosquera Pérez^1^, Jacobo Pardo-Seco^1,2^, Lidia Piñeiro Rodríguez^1^, Sara Pischedda^1,2^, Sara Rey Vázquez^1^, Irene Rivero Calle^1^, Carmen Rodríguez-Tenreiro^1^, Lorenzo Redondo-Collazo^1^, Miguel Sadiki Ora^1^, Antonio Salas^1,2^, Sonia Serén Fernández^1^, Cristina Serén Trasorras^1^, Marisol Vilas Iglesias^1^.

^1^ Translational Pediatrics and Infectious Diseases, Pediatrics Department, Hospital Clínico Universitario de Santiago, Santiago de Compostela, Spain, and GENVIP Research Group (www.genvip.org), Instituto de Investigación Sanitaria de Santiago, Universidad de Santiago de Compostela, Galicia, Spain.

^2^ Unidade de Xenética, Departamento de Anatomía Patolóxica e Ciencias Forenses, Instituto de Ciencias Forenses, Facultade de Medicina, Universidade de Santiago de Compostela, and GenPop Research Group, Instituto de Investigaciones Sanitarias (IDIS), Hospital Clínico Universitario de Santiago, Galicia, Spain

^3^ Fundación Pública Galega de Medicina Xenómica, Servizo Galego de Saúde (SERGAS), Instituto de Investigaciones Sanitarias (IDIS), and Grupo de Medicina Xenómica, Centro de Investigación Biomédica en Red de Enfermedades Raras (CIBERER), Universidade de Santiago de Compostela (USC), Santiago de Compostela, Spain

**PARTNER: RSU (Latvia)**

Principal Investigator

Dace Zavadska^1,2^

Other RSU group authors (in alphabetical order):

Anda Balode^1,2^, Arta Bārzdiņa^1,2^, Dārta Deksne^1,2^, Dace Gardovska^1,2^, Dagne Grāvele^2^, Ilze Grope^1,2^, Anija Meiere^1,2^, Ieva Nokalna^1,2^, Jana Pavāre^1,2^, Zanda Pučuka^1,2^, Katrīna Selecka^1,2^, Aleksandra Sidorova^1,2^, Dace Svile^2^, Urzula Nora Urbāne^1,2^.

^1^ Riga Stradins university, Riga, Latvia.

^2^ Children clinical university hospital, Riga, Latvia.

**PARTNER: Medical Research Council Unit The Gambia (MRCG) at LSHTM**

Principal Investigator

Effua Usuf

Additional Investigators

Kalifa Bojang

Syed M. A. Zaman

Fatou Secka

Suzanne Anderson

Anna RocaIsatou Sarr

Momodou Saidykhan

Saffiatou Darboe

Samba Ceesay

Umberto D’alessandro

Medical Research Council Unit The Gambia at LSHTM

P O Box 273,

Fajara, The Gambia

**PARTNER: ERASMUS MC-Sophia Children’s Hospital (Netherlands**

Principal Investigator

Henriëtte A. Moll¹

Research Group (alphabetical order)

Dorine M. Borensztajn¹, Nienke N. Hagedoorn, Chantal Tan ¹, ¹, Clementien L. Vermont², Joany Zachariasse ¹

Additional investigator

W Dik ^3^

¹ Erasmus MC-Sophia Children’s Hospital, Department of General Paediatrics, Rotterdam, the Netherlands

² Erasmus MC-Sophia Children’s Hospital, Department of Paediatric Infectious Diseases & Immunology, Rotterdam, the Netherlands

^3^ Erasmus MC, Department of immunology, Rotterdam, the Netherlands

**PARTNER: Swiss Pediatric Sepsis Study (Switzerland)**

Principal Investigators*:*

Philipp Agyeman, MD ^1^ (ORCID 0000-0002-8339-5444), Luregn J Schlapbach, MD, FCICM ^2,3^ (ORCID 0000-0003-2281-2598)

Clinical recruitment at University Children’s Hospital Bern for PERFORM:

Christoph Aebi ^1^, Verena Wyss ^1^, Mariama Usman ^1^

Principal and co-investigators for the Swiss Pediatric Sepsis Study:

Philipp Agyeman, MD ^1^, Luregn J Schlapbach, MD, FCICM ^2,3^, Eric Giannoni, MD ^4,5^, Martin Stocker, MD ^6^, Klara M Posfay-Barbe, MD ^7^, Ulrich Heininger, MD ^8^, Sara Bernhard-Stirnemann, MD ^9^, Anita Niederer-Loher, MD ^10^, Christian Kahlert, MD ^10^, Giancarlo Natalucci, MD ^11^, Christa Relly, MD ^12^, Thomas Riedel, MD ^13^, Christoph Aebi, MD ^1^, Christoph Berger, MD ^12^ for the Swiss Pediatric Sepsis Study

^1^ Department of Pediatrics, Inselspital, Bern University Hospital, University of Bern, Switzerland

^2^ Neonatal and Pediatric Intensive Care Unit, Children’s Research Center, University Children’s Hospital Zurich, University of Zurich, Zurich, Switzerland

^3^Child Health Research Centre, University of Queensland, and Queensland Children`s Hospital, Brisbane, Australia

^4^ Clinic of Neonatology, Department Mother-Woman-Child, Lausanne University Hospital and University of Lausanne, Switzerland

^5^ Infectious Diseases Service, Department of Medicine, Lausanne University Hospital and University of Lausanne, Switzerland

^6^ Department of Pediatrics, Children’s Hospital Lucerne, Lucerne, Switzerland

^7^ Pediatric Infectious Diseases Unit, Children’s Hospital of Geneva, University Hospitals of Geneva, Geneva, Switzerland

^8^ Infectious Diseases and Vaccinology, University of Basel Children’s Hospital, Basel, Switzerland

^9^ Children’s Hospital Aarau, Aarau, Switzerland

^10^ Division of Infectious Diseases and Hospital Epidemiology, Children’s Hospital of Eastern Switzerland St. Gallen, St. Gallen, Switzerland

^11^ Department of Neonatology, University Hospital Zurich, Zurich, Switzerland

^12^ Division of Infectious Diseases and Hospital Epidemiology, and Children’s Research Center, University Children’s Hospital Zurich, Switzerland

^13^ Children’s Hospital Chur, Chur, Switzerland

**PARTNER: Liverpool (UK)**

Principal Investigators

Enitan D Carrol^1,2,3^

Stéphane Paulus ^1,^

Research Group (alphabetical order)

Elizabeth Cocklin^1^, Rebecca Jennings^4^, Joanne Johnston^4^, Simon Leigh^1^, Karen Newall^4^, Sam Romaine^1^

^1^ Department of Clinical Infection, Microbiology and Immunology, University of Liverpool Institute of Infection and Global Health , Liverpool, England

^2^ Alder Hey Children’s Hospital, Department of Infectious Diseases, Eaton Road, Liverpool, L12 2AP

^3^ Liverpool Health Partners, 1st Floor, Liverpool Science Park, 131 Mount Pleasant, Liverpool, L3 5TF

^4^Alder Hey Children’s Hospital, Clinical Research Business Unit, Eaton Road, Liverpool, L12 2AP

**PARTNER: NKUA (Greece)**

Principal investigator

Professor Maria Tsolia (all activities)

Investigator/Research fellow

Irini Eleftheriou (all activities)

Additional investigators

Recruitment: Maria Tambouratzi

Lab: Antonis Marmarinos (Quality Manager)

Lab: Marietta Xagorari

Kelly Syggelou

2nd Department of Pediatrics, National and Kapodistrian University of Athens,

“P. and A. Kyriakou” Children’s Hospital

Thivon and Levadias

Goudi, Athens

**PARTNER: Micropathology Ltd (UK)**

Principal Investigator

Professor Colin Fink^1^, Clinical Microbiologist

Additional investigators

Dr Marie Voice^1^, Post doc scientist

Dr. Leo Calvo-Bado^1^, Post doc scientist

^1^ Micropathology Ltd, The Venture Center, University of Warwick Science Park, Sir William Lyons Road, Coventry, CV4 7EZ.

**PARTNER: Medical University of Graz (MUG, Austria)**

Principal Investigator

Werner Zenz^1^ (all activities)

Co-investigators (alphabetical order)

Benno Kohlmaier^1^ (all activities)

Nina A. Schweintzger^1^ (all activities)

Manfred G. Sagmeister^1^ (study design, consortium wide sample management)

Research team

Daniela S. Kohlfürst^1^ (study design)

Christoph Zurl^1^ (BIVA PIC)

Alexander Binder^1^ (grant application)

Recruitment team, data managers, (alphabetical order)

Susanne Hösele^1^, Manuel Leitner^1^, Lena Pölz^1^, Glorija Rajic^1^,

Clinical recruitment partners (alphabetical order)

Sebastian Bauchinger^1^, Hinrich Baumgart^4^, Martin Benesch^3^, Astrid Ceolotto^1^, Ernst Eber^2^, Siegfried Gallistl^1^, Gunther Gores^5^, Harald Haidl^1^, Almuthe Hauer^1^, Christa Hude^1^, Markus Keldorfer^5^, Larissa Krenn^4^, Heidemarie Pilch^5^, Andreas Pfleger^2^, Klaus Pfurtscheller^4^, Gudrun Nordberg^5^, Tobias Niedrist^8^, Siegfried Rödl^4^, Andrea Skrabl-Baumgartner^1^, Matthias Sperl^7^, Laura Stampfer^5^, Volker Strenger^3^, Holger Till^6^, Andreas Trobisch^5^, Sabine Löffler^5^

^1^ Department of Pediatrics and Adolescent Medicine, Division of General Pediatrics, Medical University of Graz, Graz, Austria

^2^Department of Pediatric Pulmonology, Medical University of Graz, Graz, Austria

^3^Department of Pediatric Hematooncoloy, Medical University of Graz, Graz, Austria

^4^Paediatric Intensive Care Unit, Medical University of Graz, Graz, Austria

^5^University Clinic of Paediatrics and Adolescent Medicine Graz, Medical University Graz, Graz,Austria

^6^Department of Paediatric and Adolescence Surgery, Medical University Graz, Graz, Austria

^7^Department of Pediatric Orthopedics, Medical University Graz, Graz, Austria

^8^Clinical Institute of Medical and Chemical Laboratory Diagnostics, Medical University Graz, Graz, Austria

**PARTNER: London School of Hygiene and Tropical Medicine (UK)**

WP 1 WP2, WP5

Principal Investigator:

Dr Shunmay Yeung^1,2 3^ PhD, MBBS, FRCPCH, MRCP, DTM&H

Research Group

Dr Juan Emmanuel Dewez^1^ MD, DTM&H, MSc

Prof Martin Hibberd ^1^ BSc, PhD

Mr David Bath^2^ MSc, MAppFin, BA(Hons)

Dr Alec Miners^2^ BA(Hons), MSc, PhD

Dr Ruud Nijman^3^ PhD MSc MD MRCPCH

Dr Catherine Wedderburn^1^ BA, MBChB, DTM&H, MSc, MRCPCH

Ms Anne Meierford^1^ MSc, BMedSc, BMBS

Dr Baptiste Leurent^4^, PhD, MSc

1. Faculty of Infectious and Tropical Disease, London School of Hygiene and Tropical Medicine, London, UK
2. Faculty of Public Health and Policy, London School of Hygiene and Tropical Medicine, London, UK
3. Department of Paediatrics, St. Mary’s Hospital Imperial College Hospital, London, UK
4. Faculty of Epidemiology and Population Health, London School of Hygiene and Tropical Medicine, London, UK

**PARTNER: Radboud University Medical Center (RUMC, Netherlands)**

Principal Investigators

Ronald de Groot ^1^, Michiel van der Flier ^1,2,3^, Marien I. de Jonge^1^

Co-investigators Radboud University Medical Center (alphabetical order)

Koen van Aerde^1,2^, Wynand Alkema^1^, Bryan van den Broek^1^, Jolein Gloerich^1^, Alain J. van Gool^1^, Stefanie Henriet^1,2^, Martijn Huijnen^1^, Ria Philipsen^1^, Esther Willems^1^

Investigators PeDBIG PERFORM DUTCH CLINICAL NETWORK (alphabetical order)

G.P.J.M. Gerrits^8^, M. van Leur^8,^ J. Heidema ^4^,L. de Haan^1,2^ C.J. Miedema ^5^, C. Neeleman ^1^ C.C. Obihara ^6^, G.A. Tramper-Stranders7^6^

1. Radboud University Medical Center, Nijmegen, The Netherlands
2. Amalia Children’s Hospital, Nijmegen, The Netherlands
3. Wilhelmina Children’s Hospital, University Medical Center Utrecht, Utrecht, The Netherlands
4. St. Antonius Hospital, Nieuwegein, The Netherlands
5. Catharina Hospital, Eindhoven, The Netherlands
6. ETZ Elisabeth, Tilburg, The Netherlands
7. Franciscus Gasthuis, Rotterdam, The Netherlands
8. Canisius Wilhelmina Hospital, Nijmegen, The Netherlands

**PARTNER: Oxford (UK)**

Principal Investigators

Andrew J. Pollard^1,2^, Rama Kandasamy^1,2^, Stéphane Paulus ^1,2^

Additional Investigators

Michael J. Carter^1,2^, Daniel O'Connor^1,2^, Sagida Bibi^1,2^, Dominic F. Kelly^1,2^, Meeru Gurung^3^, Stephen Thorson^3^, Imran Ansari^3^, David R. Murdoch^4^, Shrijana Shrestha^3^.

^1^Oxford Vaccine Group, Department of Paediatrics, University of Oxford, Oxford, United Kingdom.

^2^NIHR Oxford Biomedical Research Centre, Oxford, United Kingdom.

^3^Paediatric Research Unit, Patan Academy of Health Sciences, Kathmandu, Nepal.

^4^Department of Pathology, University of Otago, Christchurch, New Zealand.

**PARTNER: Newcastle University, Newcastle upon Tyne, (UK)**

Principal Investigator

Marieke Emonts ^1,2,3^ (all activities)

Co-investigators

Emma Lim^2,3,7^ (all activities)

Lucille Valentine^4^

Recruitment team (alphabetical), data-managers, and GNCH Research unit

Karen Allen^5^, Kathryn Bell^5^, Adora Chan^5^, Stephen Crulley^5^, Kirsty Devine^5^, Daniel Fabian^5^, Sharon King^5^, Paul McAlinden^5^, Sam McDonald^5^, Anne McDonnell2,^5^, Ailsa Pickering^2,5^, Evelyn Thomson^5^, Amanda Wood^5^, Diane Wallia^5^, Phil Woodsford^5^,

Sample processing: Frances Baxter^5^, Ashley Bell^5^, Mathew Rhodes^5^

PICU recruitment

Rachel Agbeko^8^

Christine Mackerness^8^

Students MOFICHE

Bryan Baas^2^, Lieke Kloosterhuis^2^, Wilma Oosthoek^2^

Students/medical staff PERFORM

Tasnim Arif^6^, Joshua Bennet^2^, Kalvin Collings^2^, Ilona van der Giessen^2^, Alex Martin^2^, Aqeela Rashid^6^, Emily Rowlands^2^, Gabriella de Vries^2^, Fabian van der Velden^2^

Engagement work/ethics/cost effectiveness

Lucille Valentine ^4^, Mike Martin^9^, Ravi Mistry^2^, Lucille Valentine^4^

^1^ Translational and Clinical Research Institute, Newcastle University, Newcastle upon Tyne UK

^2^Great North Children’s Hospital, Paediatric Immunology, Infectious Diseases & Allergy, Newcastle upon Tyne Hospitals NHS Foundation Trust, Newcastle upon Tyne, United Kingdom.

^3^NIHR Newcastle Biomedical Research Centre based at Newcastle upon Tyne Hospitals NHS Trust and Newcastle University, Westgate Rd, Newcastle upon Tyne NE4 5PL, United Kingdom

^4^Newcastle University Business School, Centre for Knowledge, Innovation, Technology and Enterprise (KITE), Newcastle upon Tyne, United Kingdom

^5^Great North Children’s Hospital, Research Unit, Newcastle upon Tyne Hospitals NHS Foundation Trust, Newcastle upon Tyne, United Kingdom.

^6^Great North Children’s Hospital, Paediatric Oncology, Newcastle upon Tyne Hospitals NHS Foundation Trust, Newcastle upon Tyne, United Kingdom.

^7^Population Health Sciences Institute, Newcastle University, Newcastle upon Tyne, UK

^8^Great North Children’s Hospital, Paediatric Intensive Care Unit, Newcastle upon Tyne Hospitals NHS Foundation Trust, Newcastle upon Tyne, United Kingdom.

^9^Northumbria University, Newcastle upon Tyne, United Kingdom.

**PARTNER: LMU Munich (Germany)**

Principal Investigator

Ulrich von Both^1,2^ MD, FRCPCH (all activities)

Research group

Laura Kolberg¹ MSc (all activities)

Manuela Zwerenz¹ MSc, Judith Buschbeck¹ PhD

Clinical recruitment partners (alphabetical order)

Christoph Bidlingmaier^3^, Vera Binder^4^, Katharina Danhauser^5^, Nikolaus Haas^10^, Matthias Griese^6^, Tobias Feuchtinger^4^, Julia Keil^9^, Matthias Kappler^6^, Eberhard Lurz^7^, Georg Muench^8^, Karl Reiter^9^, Carola Schoen^9^

¹Div. Paediatric Infectious Diseases, Hauner Children’s Hospital, University Hospital, Ludwig Maximilians University (LMU), Munich, Germany

^2^German Center for Infection Research (DZIF), Partner Site Munich, Munich, Germany

^3^Div. of General Paediatrics, ^4^Div. Paediatric Haematology & Oncology, ^5^Div. of Paediatric Rheumatology, ^6^Div. of Paediatric Pulmonology, ^7^Div. of Paediatric Gastroenterology, ^8^Neonatal Intensive Care Unit, ^9^Paediatric Intensive Care Unit Hauner Children’s Hospital, University Hospital, Ludwig Maximilians University (LMU), Munich, Germany, ^10^Department Pediatric Cardiology and Pediatric Intensive Care, University Hospital, Ludwig Maximilians University (LMU), Munich, Germany

**PARTNER: bioMérieux (France)**

Principal Investigator

François Mallet^1,2, 3^

Research Group

Karen Brengel-Pesce^1,2, 3^

Alexandre Pachot^1^

Marine Mommert^1,2^

^1^Open Innovation & Partnerships (OIP), bioMérieux S.A., Marcy l'Etoile, France

^2^ Joint research unit Hospice Civils de Lyon - bioMérieux, Centre Hospitalier Lyon Sud, 165 Chemin du Grand Revoyet, 69310 Pierre-Bénite, France

^3^ EA 7426 Pathophysiology of Injury-induced Immunosuppression, University of Lyon1-Hospices Civils de Lyon-bioMérieux, Hôpital Edouard Herriot, 5 Place d’Arsonval, 69437 Lyon Cedex 3, France

**PARTNER: University Medical Centre Ljubljana (Slovenia)**

Principal Investigator

Marko Pokorn^1,2,3^ MD, PhD

Research Group

Mojca Kolnik^1^ MD, Katarina Vincek^1^ MD, Tina Plankar Srovin^1^ MD, PhD, Natalija Bahovec^1^ MD, Petra Prunk^1^ MD, Veronika Osterman^1^ MD, Tanja Avramoska^1^ MD

^1^Department of Infectious Diseases, University Medical Centre Ljubljana, Japljeva 2, SI-1525 Ljubljana, Slovenia

^2^University Childrens' Hospital, University Medical Centre Ljubljana, Ljubljana, Slovenia

^3^Department of Infectious Diseases and Epidemiology, Faculty of Medicine, University of Ljubljana, Slovenia

**PARTNER: Amsterdam, Academic Medical Hospital & Sanquin Research Institute (Netherlands)**

Principal Investigator

Taco Kuijpers ^1,2^

Co-investigators

Ilse Jongerius ^2^

Recruitment team (EUCLIDS, PERFORM)

J.M. van den Berg^1^, D. Schonenberg^1^, A.M. Barendregt^1^, D. Pajkrt^1^, M. van der Kuip^1,3^, A.M. van Furth^1,3^

Students PERFORM

Evelien Sprenkeler ^2^, Judith Zandstra ^2^

Technical support PERFORM

G. van Mierlo ^2^, J. Geissler ^2^

^1^ Amsterdam University Medical Center (Amsterdam UMC), location Academic Medical Center (AMC), Dept of Pediatric Immunology, Rheumatology and Infectious Diseases, University of Amsterdam, Amsterdam, the Netherlands

^2^ Sanquin Research Institute, & Landsteiner Laboratory at the AMC, University of Amsterdam, Amsterdam, the Netherlands.

^3^ Amsterdam University Medical Center (Amsterdam UMC), location Vrije Universiteit Medical Center (VUMC), Dept of Pediatric Infectious Diseases and Immunology, Free University (VU), Amsterdam, the Netherlands (former affiliation)
